# Impact of COVID-19 on Care-Home Mortality and Life Expectancy in Scotland

**DOI:** 10.1101/2021.01.15.21249871

**Authors:** Jennifer K Burton, Martin Reid, Ciara Gribben, David Caldwell, David N Clark, Peter Hanlon, Terence J Quinn, Colin Fischbacher, Peter Knight, Bruce Guthrie, David A McAllister

## Abstract

**Introduction:** COVID-19 deaths are commoner among care-home residents, but the mortality burden has not been quantified.

**Methods:** Care-home residency was identified via a national primary care registration database linked to national mortality data. Life expectancy was estimated using Makeham-Gompertz models, to (i) describe yearly life expectancy from Nov 2015 to Oct 2020 (ii) compare life expectancy (during 2016-2018) between care-home residents and the wider Scottish population and (iii) apply care-home life expectancy estimates to COVID-19 death counts to estimate years of life lost (YLL).

**Results:** Among care-home residents, life expectancy in 2015/16 to 2019/20 ranged from 2.7 to 2.3 years for women and 2.3 to 1.8 years for men. Life expectancy was lowest in 2019/20. Age-sex specific life expectancy in 2016-2018 in care-home residents was lower than in the Scottish population (10 and 2.5 years in those aged 70 and 90 respectively). Rather than using national life tables, applying care-home specific life expectancies to COVID-19 deaths yields, mean YLLs for care-home residents were 2.6 and 2.2 for women and men respectively, with total care-home resident YLLs of 3,560 years in women and 2,046 years in men. In people aged over-70, approximately half of deaths and a quarter of YLL attributed to COVID-19 were accounted for by the 5% of over-70s who were care-home residents.

**Conclusion:** Prioritising care-home residents for vaccination is justified not only in terms of total deaths, but also in terms of years of life lost.

**Research in context:** *Evidence before this study:* We searched PubMed to 1^st^ December 2020, with the terms (“nursing home” OR “care-home” OR “long-term care” OR “residential care”) AND (“mortality” OR “life expectancy” OR “length of stay”). We also searched for studies specific to the impact of the COVID-19 pandemic on those living in care-homes. We restricted our search to publications in English. Usual care-home life expectancy, in a UK context, has not previously been defined. One systematic review of length of stay was identified, which found significant heterogeneity in factors and associations. The impact of COVID-19 on excess mortality among care-home residents was noted, but the impact on life expectancy was not reported. Studies evaluating life expectancy among older people in the COVID-19 pandemic have not taken account of residency in their estimates.

*Added value of this study:* Using Scottish national representative linked data we describe the usual life expectancy of older adults (aged ≥70 years) living in care-homes, compared to older people living elsewhere. Deaths among care-home residents account for a considerable proportion of all mortality in older adults, around 19% for men and 30% for women. Life expectancy in care-home residents during the pandemic fell by almost 6 months, from 2.7 to 2.3 years in men and 2.1 to 1.8 years in women. In total, over 5,600 Years of Life were Lost (YLL) by care-home residents in Scotland who died with COVID-19. Around half of COVID-19 deaths and a quarter of YLL in those aged 70 years and over occurred among care-home residents. During the COVID-19 pandemic a smaller proportion of deaths among care-home residents occurred in hospitals.

*Implications of all the available evidence:* Prioritising the 5% of older adults who are care-home residents for vaccination against COVID-19 is justified both in terms of total deaths and total years of life lost. Individual and societal planning for care needs in older age relies on understanding usual care-home life expectancy and patterns of mortality. Understanding life expectancy may help clinicians, residents and their families make decisions about their health care, facilitating more informed discussions around their priorities and wishes. Population-wide estimates of YLL and burden of disease should take account of residency status, given the significant differences between life expectancy of those living in care-homes from their peers in other settings.

## Background

COVID-19 has severely impacted people living in care-homes.^1,2^ Residents have experienced significant morbidity and mortality,^3,4^ but also reduced access to routine healthcare and other services, and disconnection from family and friends.^5^ Striking the correct balance between minimising loss of life from COVID-19 and maintaining the quality of life of care-home residents has proved controversial.^6,7^

Care-homes in Scotland are home to around 5% of the population aged over-70 years.^8,9^ Compared to older people in the general population, people living in care-homes in the UK are known to have higher mortality.^10-12^ Care-homes largely care for people approaching the end of their lives,^13^ so higher mortality is expected, but there is limited contemporary population-level data on the life expectancy of care-home residents.^14^ Moreover, while the life expectancy (i.e. the notional years of life lost) of people dying with COVID-19 in the community has been estimated,^15,16^ there is no published data on the years of life lost (YLL) among care-home residents who died with COVID-19.

Such information is difficult to apply to individuals (who may live for a considerably longer or shorter duration than their life expectancy), but it is crucial for designing vaccination and other preventive strategies that minimise the total years of life lost across the population. Prior to the (imminent) availability of effective vaccines, COVID-19 care-home policies involved trading-off short term restrictions to quality of life against the prevention of COVID-19 morbidity and mortality. However, these trade-offs will be different in someone near the end of life compared to someone with a longer life expectancy. Indeed, this trade-off applies not only to COVID-19, but also to many drug therapies aimed at extending life rather than treating symptoms. For all these purposes, decision-making by care-home residents, their families and policymakers, requires information on the life expectancy of care-home residents.

Due to high quality routinely collected data on care-home residency status, it is possible in Scotland to address this need. Using these data, for care-home residents and other older people, we describe 5-year trends in age and sex-specific mortality, trends in life expectancy among care-home residents, and estimate the mortality impact of COVID-19 including years of life lost up to November 2020.

## Methods

### Design, population and data sources

A population-wide dynamic cohort study was created using the Community Health Index (CHI) register, based on the CHI number (a unique identifier variable used across NHS services in Scotland). All adults aged 70 years and over, registered with a GP in Scotland were included. Residency was categorised into care-home or not, based on the presence of the institution flag on records of those living at care-home addresses, applied at time of GP registration or address change. Data for care-home residents were linked to the National Records of Scotland (NRS) mortality data, which records all deaths certified in Scotland in any location. In addition, for 2020, cases of COVID-19 where an individual died were collected using data from the REACT-SCOT population-wide case-control study.^17^

### Exposures

A care-home in Scotland is defined as a facility providing 24-hour care to its residents with no regulatory distinction between ‘residential’ and ‘nursing’ homes.^18^ While some care-homes provide care for younger adults (or example, people with learning disability),^19^ we restricted this analysis to the older adult population aged 70 years and over.

Care-home residency was defined as the presence of a relevant Institution Code on the CHI register, which was available from 2007. Individuals were defined as care-home residents if they had a first CHI-flag indicating care-home residency on or after 8 years prior to each month. Eight years was chosen so that the “look-back” period for earliest month fell after the recording of care-home had been established within the CHI database. This analysis was focused on long-stay residents in care-homes. This method does not capture temporary stays for respite and intermediate care (where CHI registered address does not change).

We examined the accuracy of the CHI Institution flag, against a new measure of care-home status which was created in 2020 in response to the COVID-19 pandemic and is thus only available for 2020. A detailed description of the care-home status measure is provided in the supplementary appendix. Briefly, the Unique Property Reference Number (UPRN), an Ordnance Survey product which uniquely identifies each property in the UK, was assigned to all addresses in the CHI register for all individuals (>5.5 million) alive at any time during 2020. Each UPRN was then mapped to a definitive list of addresses of registered care-home services held by the Care Inspectorate.^9^ Using the control arm of the REACT case-control study (which is an age, sex and general practice area stratified random sample of all individuals in Scotland), we then calculated the sensitivity and specificity of the CHI care-home flag as a measure of care-home residency status on the 1^st^ of August 2020 against the new care-home status measure.

### Outcomes

Mortality was examined for all care-home residents using NRS mortality data. Causes of death were grouped based on the underlying cause of death derived from the death certificate into the following categories:-respiratory (including pneumonia) [ICD1-J00 to J99 and COVID-19 – U07), dementia [F01, F03, G30], circulatory [I00 to I97], cancer [C00 to C97], and other. Deaths in hospital were identified from the location of death recorded on the death certificate.

Life expectancy estimates for the general population were obtained from published NRS national life tables,^20^ held on the Human Mortality Database.^21^

For the years of life lost calculations for death with COVID-19, we included all deaths within 28 days of a positive SARS-CoV-2 PCR test result and/or with COVID-19 recorded as a cause of their death (underlying or otherwise). Counts of deaths with COVID-19 by age, sex and care-home residency for all individuals aged 70 years and over in Scotland were obtained from the REACT case-control study.

### Covariates

Individuals were categorised by age, sex among care-home residents and the general Scottish population. For those who died, age at death and year of death were calculated. Age at care-home admission was calculated based on date of birth and the date that the institution code was first applied to the CHI register. This was to allow inclusion of those who move between care-home services but remain resident in a care-home over several years.

### Statistical analysis

The statistical analysis was conducted in two stages. First for care home residents individual-level data was aggregated. For each calendar month from October 2015 to November 2020 we summed events and person-time by sex among care-home residents aged 70 or older at the time of death. We summed person-time and deaths overall, by cause and where the place of death was a hospital. Person time was calculated similarly as the number of days in the month minus any days prior to an individual being admitted to a care-home and any days after the individual had died. We also obtained aggregated data on deaths for all individuals aged 70 years or older in Scotland up to September 2020. Events and person-time were aggregated (i) by sex and calendar month for all years, (ii) by age, sex and each 12-months running from November 2015 to October 2020, and (iii) by age, sex and calendar year from 2016 to 2018 inclusive. The former period was chosen as this was the latest data available for care-homes, the latter period was chosen to allow comparison with published life tables from the National Records for Scotland.

Secondly, using the MortalityLaws package in R (version 2.0, R Core team, Austria), Makeham-Gompertz models were fitted to the age and sex-specific mortality rates to obtain age and sex-specific life expectancy estimates for each year and for the 3-year period from 2016-2018. Uncertainty was propagated to the estimates by fitting Makeham-Gompertz models for 1000 samples from Binomial distributions independently sampled from each stratum (defined by the age, sex, time period and care-home residency status) where the probability parameter was the number of events divided by the population at risk at the onset of each time period to obtain a distribution of estimates, and summarised via the 2.5^th^ and 97.5^th^ percentiles. For the 2016-2018 period, life expectancy was also calculated non-parametrically using life tables, and both analyses were compared to the NRS calculated life expectancy (which are also based on life tables) for the entire Scottish population.

The care-home resident and NRS age-sex specific life expectancy estimates were applied to the age-sex distribution of individuals who died with COVID-19 (from the REACT case-control study) to calculate the years of life lost among people aged 70 years and over. The years of life lost due to COVID-19, ignoring care-home residency (i.e. based solely on the NRS population life tables) and accounting for care-home residency (based on the life expectancy estimates described above) were then calculated as the age-sex specific remaining life expectancy for each individual who had died.

### Data governance

Standard Public Health Scotland policies were followed in conducting analyses using data held within PHS for the purposes of monitoring population health and supporting policymaking.

## Results

In 2017, 26,056 women and 11,086 men aged 70 years and over were resident in care-homes, representing 6% and 4% of the whole population in this age group. Of these 25% had resided in a care-home for <12 months with a further 25% for less than 24 months with a median stay of 1.85 years for men and 2.12 years for women (see supplementary appendix for full distribution).

### Validation of CHI flag

The sensitivity and specificity of the CHI care-home flag compared to the UPRN care-home flag in 2020 was excellent (96%), with only minor variation across the NHS Board regions.

### Trends in mortality and life expectancy

Deaths among care-home residents in all years 2015/16-2019/20 accounted for a considerable proportion of all mortality in older adults, around 19% for men and 30% for women. Differences between 2020 and previous years were seen in April and May. In the month of April during the years 2016-2019, 29 to 32% of deaths in women were among care-home residents, while in April 2020 the equivalent figure was 42%. Differences of similar magnitude were seen in May among women and in April and May among men. In 2019/20 13% of female and 18% of male care-home resident deaths took place in hospital, lower than the 24-25% and 16-19% in 2015/16 to 2018/19 (Table 1).

**Table 1:** Care-home resident deaths.

| Sex | November to October | % of all deaths of those aged 70 years and older which were deaths of care-home residents | % of care-home residents' deaths which occurred in hospital |
| --- | --- | --- | --- |
| Men | 2015/16 | 3470/18227 (19%) | 865/3470 (25%) |
|  | 2016/17 | 3355/19068 (18%) | 820/3355 (24%) |
|  | 2017/18 | 3575/19841 (18%) | 849/3575 (24%) |
|  | 2018/19 | 3525/19127 (18%) | 833/3525 (24%) |
|  | 2019/20 | 4389/19618 (22%) | 779/4389 (18%) |
| Women | 2015/16 | 6857/22534 (30%) | 1324/6857 (19%) |
|  | 2016/17 | 6748/23415 (29%) | 1180/6748 (17%) |
|  | 2017/18 | 7137/24089 (30%) | 1171/7137 (16%) |
|  | 2018/19 | 6555/22757 (29%) | 1050/6555 (16%) |
|  | 2019/20 | 7970/23364 (34%) | 1024/7970 (13%) |

For this table the period was October to September (rather than November to October which was used for the formal analyses). This was done to allow comparison with quarterly deaths data released by NRS for people aged 70 and older.

Figure 1 shows mortality in care-home residents compared to the general Scottish population. As well as the large peak in spring 2020, there was a peak in winter 2017/18 (which is known to have been a severe year for seasonal influenza)^22^ and a smaller peak in winter 2019/20.

**Figure 1:**
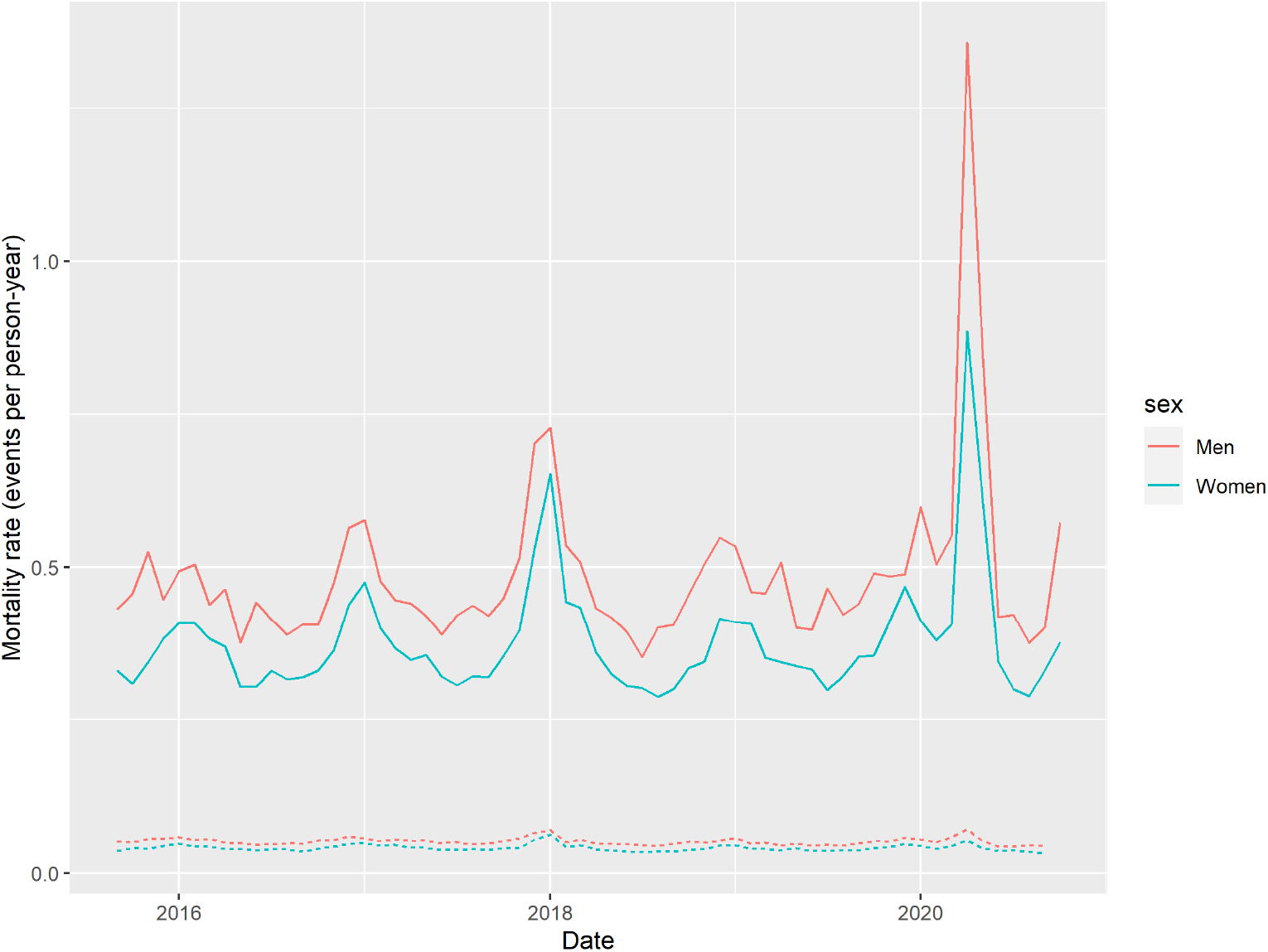
Sex-specific mortality in care-home residents compared to the general Scottish population mortality, October 2015 to November 2020. Red lines are rates in women and blue in men. The rate is per (single) person year. Solid lines indicate care-home residents aged 70 years and over and dotted lines the general population aged 70 years and over comparison.

This increased mortality in November 2019 to October 2020, and to a lesser extent the same period in 2017/18, is reflected in life expectancy estimates for these periods. Among women aged 70 years and over living in care-homes life expectancy was 2.7, 2.6, 2.5, 2.7 and 2.3 years respectively for 2015/16 to 2019/20. For men, corresponding figures were 2.3, 2.1, 2.1, 2.1 and 1.8 years. Compared to the highest year, life expectancy in care-home residents fell by around 2-2.5 months in 2017/18 and around 5.5 months in 2019/20 (Table 2, Figure 2).

**Table 2:**
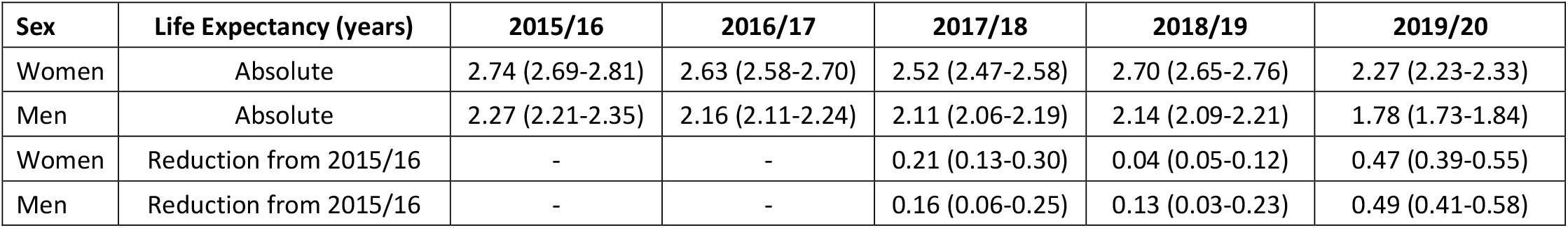
Life expectancy estimates for November to October 2015/16-2019/20 for care-home residents.

**Figure 2:**
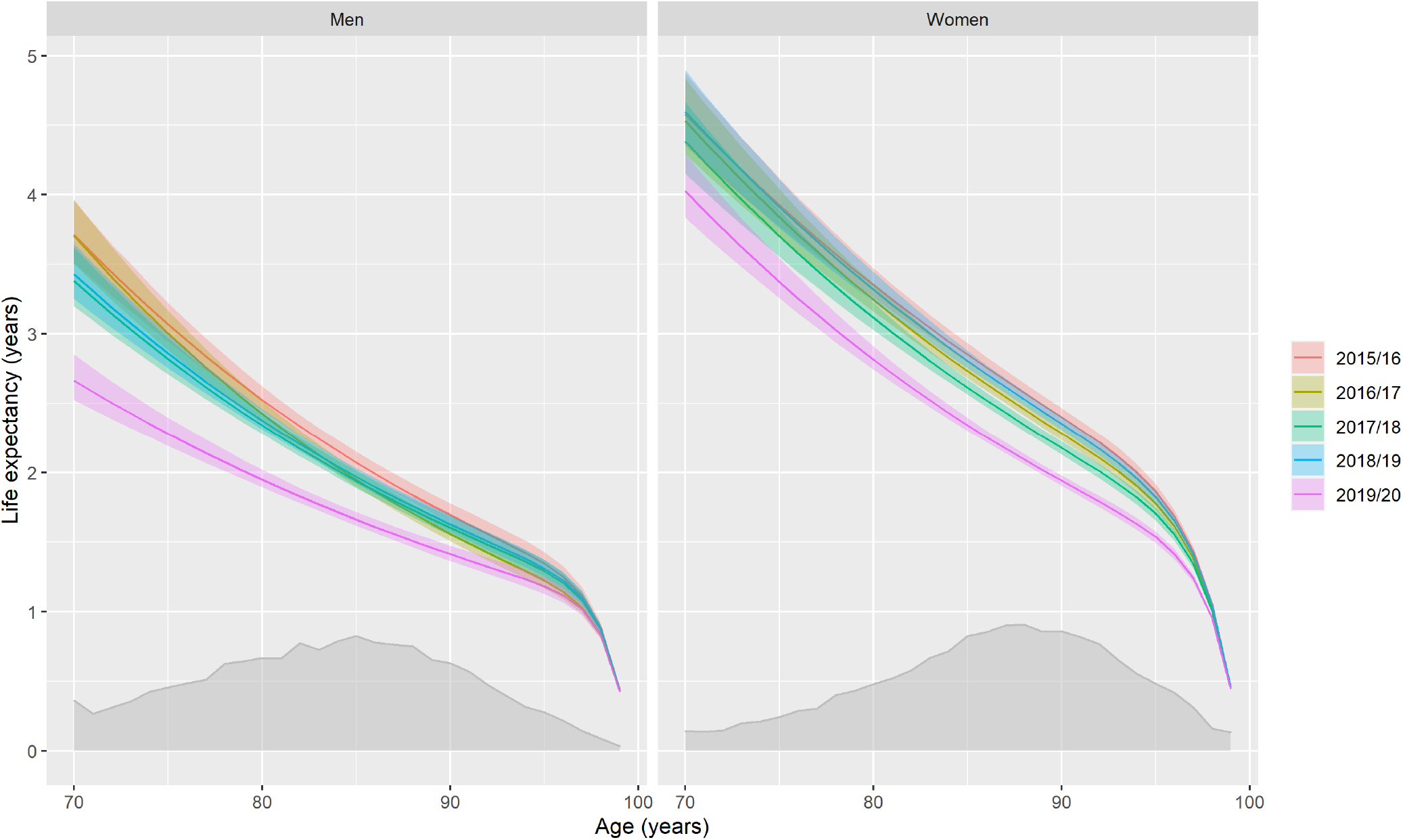
Age and sex-specific life expectancy estimates for care-home residents with uncertainty intervals, 2015/16-2019/20. Shaded area denotes the age-sex-distribution of care-home residents in 2017. Shaded area denotes the age and sex distribution of the care-home population in 2017 (to retain clarity in the figure and since absolute values of probability densities are not easily interpretable without further calculation no secondary axis is shown).

Figure 3 shows the mortality by underlying cause of death for the same period. In 2019/20 more than half of the mortality peak was deaths attributed to respiratory disease, mostly COVID-19, with an increase also seen in circulatory diseases. Dementia was the second largest single recorded cause of care-home deaths in 2019/20.

**Figure 3:**
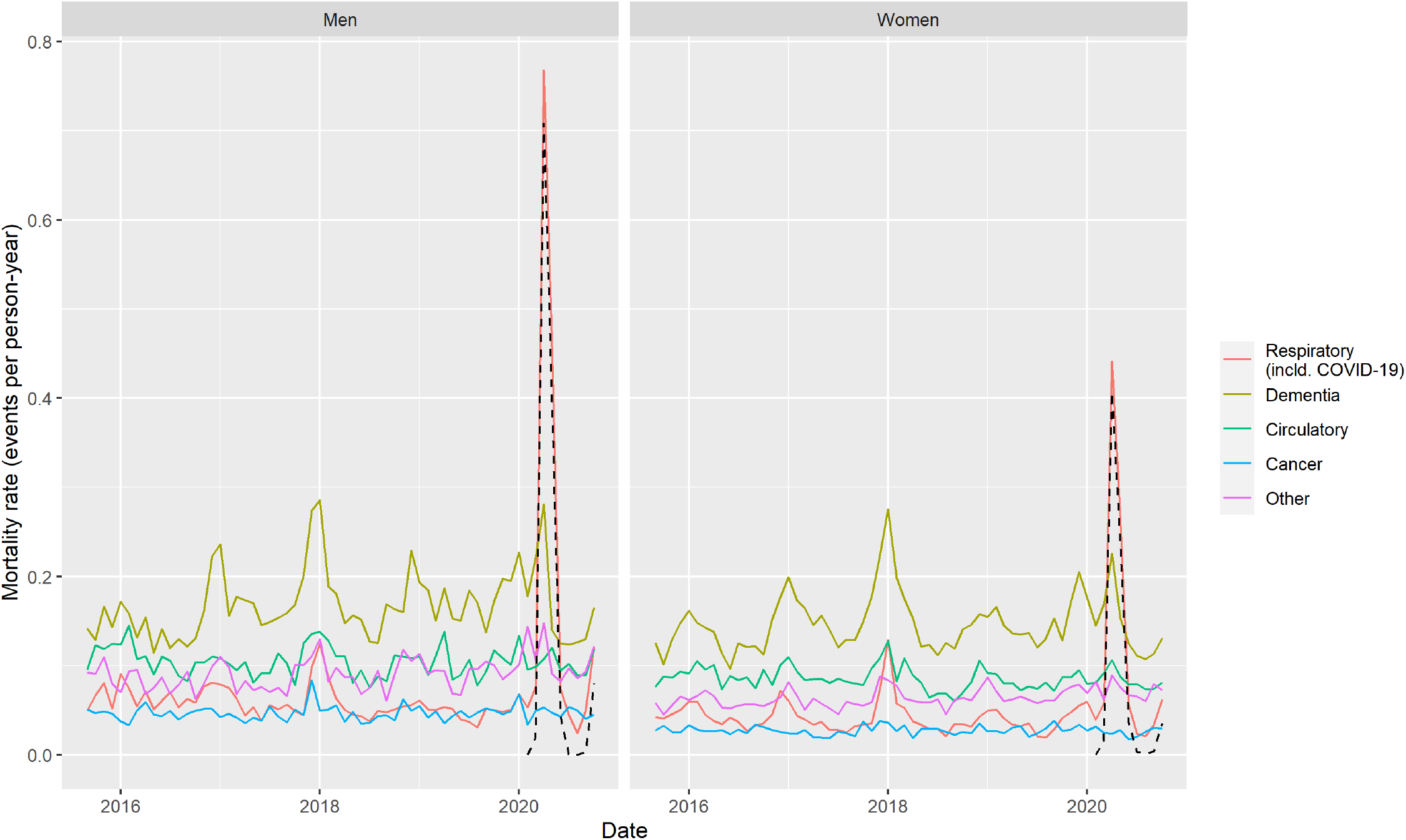
Sex-specific mortality by cause in care-home residents, October 2015 to November 2020. Dotted line indicates deaths with COVID-19 as the underlying cause which are a subset of respiratory deaths. All other causes are mutually exclusive.

### Life expectancy and years of life lost in people dying with COVID-19

Figure 4 shows the 2016-2018 age- and sex-specific life expectancy for adults aged 70 years and over. It shows both the standard total population estimates (from NRS) and estimates we produced for care-home residents in this analysis. Care-home residents have substantially shorter life expectancies than those living in non-institutional settings, with larger differences at younger ages (e.g., around 10 years aged 70 and 2.5 years aged 90). Averaging over the age-distribution of care-home residents alive in 2017, led to a difference of around 5 years.

**Figure 4:**
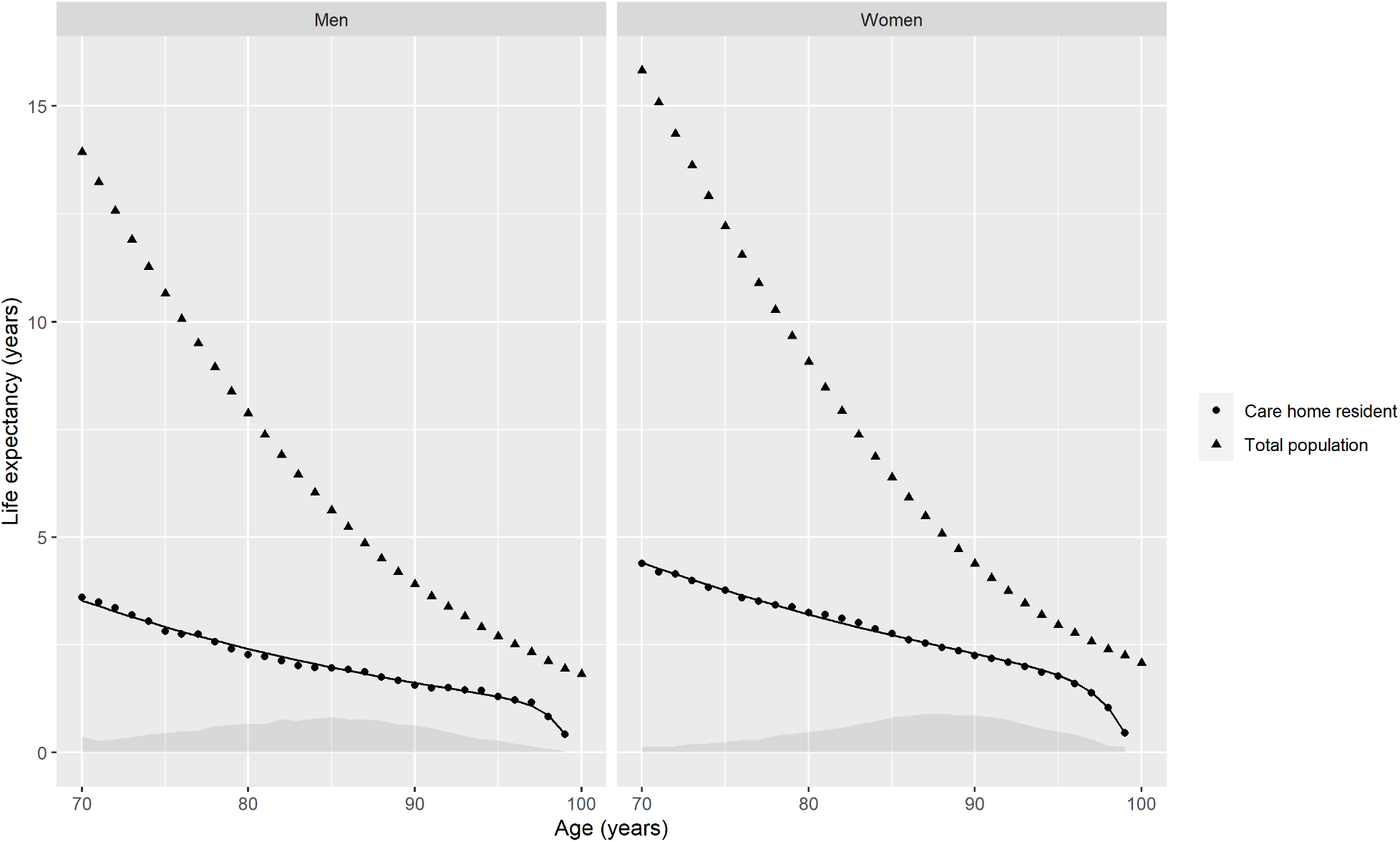
Age and sex-specific life expectancy in care-home residents compared to the age and sex specific general Scottish population, 2016-2018. Shaded area denotes the age and sex distribution of the care-home population in 2017 (to retain clarity in the figure and since absolute values of probability densities are not easily interpretable without further calculation no secondary axis is shown). Calendar year data was used here rather than a 12-month which maximised the months spent within the pandemic in order to allow comparison between the care home residents life expectancy estimates and that obtained from the NRS life tables.

Table 3 shows the estimated years of life lost (YLL) in people aged 70 years and over obtained by applying these age-sex and residency specific life expectancy estimates to the 1370 women and 943 men who were care-home residents, and the 927 women and 1204 men who were not care-home residents, who died of COVID-19. Among care-home residents, the impact of estimating YLL using care-home specific life expectancy estimates rather than general population life tables was large, leading to a difference of around 4-years; on using care-home specific life expectancy estimates for women, the estimated average YLL fell from 6.24 years per death to 2.60 years per death. For men, the equivalent figures were 6.73 and 2.17.

**Table 3:** Years of life lost among over-70s due to COVID-19 March to November 2020.

|  | Women |  | Men |  |
| --- | --- | --- | --- | --- |
| Life expectancy method used for YLL calculation | NRS solely | NRS and care-home | NRS solely | NRS and care-home |
| Care-home deaths | 1370 | 1370 | 943 | 943 |
| Not care-home deaths | 927 | 927 | 1204 | 1204 |
| Overall deaths | 2297 | 2297 | 2147 | 2147 |
| Total YLL Care-home | 8553 | 3560 | 6349 | 2046 |
| Total YLL Not care-home | 7652 | 7652 | 9520 | 9520 |
| **Total YLL Overall** | **16204** | **11211** | **15870** | **11566** |
| Average YLL Care-home | 6.24 | 2.60 | 6.73 | 2.17 |
| Average YLL Not care-home | 8.25 | 8.25 | 7.91 | 7.91 |
| **Average YLL Overall** | **7.05** | **4.88** | **7.39** | **5.39** |

*The total YLL is the sum of all YLL of those who died, and the average YLL is this number divided by the total number of deaths. “NRS solely” indicates that national life tables were used to estimate the YLL for all deaths regardless of care home residency. “NRS and care-home” indicates that care home specific life expectancy estimates were used to estimate deaths for care home residents and national life tables were used for deaths in individuals not resident in care homes. The NRS life tables are based on all deaths, including care home residents and so will slightly under-estimate LE in individuals who are not residents of care home residents.*

Care-home residents only account for approximately 5% of the Scottish population of people aged 70 years and older, but they account for 52% of all COVID-19 deaths in this age-group (and 44% of all COVID-19 deaths). Using care-home specific life expectancy estimates to calculate YLL in care-home residents, the average YLL for care-home residents remained >2 years for both men and women, corresponding to a total YLL of 3,560 years in women and 2,046 years in men. At a whole population level, recalculating YLL based on care-home specific life expectancy has a large effect on the overall YLL attributable to COVID-19. In the over-70s this fell from 7.05 years to 4.88 years in women and 7.39 years to 5.39 years in men.

## Discussion

This analysis provides the first population-level data reporting care-home life expectancy and evaluating the impact of COVID-19 on those living in care-homes in terms of YLL in the UK NHS context, and we make several important observations.

Life expectancy in care-home residents during the pandemic fell by almost 6 months. The estimated YLL due to COVID-19 is substantial for care-home residents at around two and half years per death for women and two years per death for men, corresponding to a total of around 3,600 YLL in women and 2,000 YLL in men. Although care-home residents are only ∼5% of the population aged 70 years and older, care-home resident YLL represent around a quarter of all YLL among those in that age group who died with COVID-19.

In the period prior to the pandemic life expectancy in care-home residents was substantially lower than in non-care-home residents; consequently the YLL per death is estimated to be around 4 years lower when care-home specific life expectancy rather than national life tables are used for the calculation. Since around half of COVID-19 deaths in those aged 70 years and over were among care-home residents this has a large impact at the level of the population, with the average YLL being around five years rather than seven among all those aged 70 years and over. Thus calculations of YLL for COVID-19 should use care home specific life expectancy estimates rather than standard national life tables.

Our findings are broadly consistent with studies reporting COVID-19 mortality among care-home residents in Scotland,^3^ elsewhere in the UK, ^4,23^ and internationally.^24,25^ In one UK analysis based on data (including primary care data) from Wales there was found to be excess mortality among care-home residents even after accounting for frailty.^23^ When we expressed the mortality impact in terms of life expectancy, the impact of the pandemic on mortality from December 2019 to November 2020 was more than 2-fold larger than that seen during 2017/2018, which included a severe influenza season. However, this observation needs to be seen in the context of the considerable efforts to prevent entry of SARS-CoV2 to care-homes, and the fact that only 32% of care-homes had any cases of COVID-19.^26^

Some may be surprised that the life expectancy in care-home residents was not shorter than the 2-2.5 years we observed. It is important to note that our analysis does not include individuals who died within 6-weeks or so of entering care-homes (who did not have a change of address recorded) and as such is applicable only to residents who survive beyond this initial period. Nonetheless, our findings are consistent with our previous work on a smaller sample of around 22,000 care-home resident deaths in Scotland using the Scottish Care-Home Census data over four years,^19^ and with a previous study in England and Wales which found that the one-year mortality was 26%.^10^

Our findings also have implications for studies examining the impact of COVID-19 and the pandemic response on wider society, where these rely on estimates of years of life lost or related metrics such as QALYs and DALYs. We and others have reported YLL with COVID-19^15,27^ of around 10 years in the general population. We accounted for multimorbidity,^15^ but not care-home residency. Our current findings suggest that this is an important omission; the combination of a large difference in life expectancy and a large proportion of COVID-19 deaths occurring in care-homes means that estimates which do not account for care-home residency substantially over-estimate the total YLL in the population.

### Limitations of the study

This analysis is based on Scotland-wide registration systems and is therefore highly representative. Use of anonymised, linked routinely collected data is inclusive, and not reliant on individual consent – an important consideration when analysing data on the population living in care-homes as it will lead to reduced bias.^28^ GP registration is necessary as a means of accessing primary healthcare in Scotland and thus is likely to represent a highly representative sampling frame when the focus is on individuals aged 70 years and over. Our validation work using UPRN in 2020 is reassuring in terms of the accuracy of the CHI-based identification of care-home status. Nonetheless, under-ascertainment of care-home residents is possible, though difficult to quantify. Data are published on care-home places (around 40,000 in 2020), but occupancy is not routinely collected or reported and some care-homes operate exclusively for non-resident short-stays and respite care. Previous analyses identified around 35,000 long-stay residents in Scotland’s care-homes at all ages^19^ and our data broadly align with these figures. However, we are likely to have missed those with very short care-home stays (e.g. those discharged to care-homes for end of life care who die before their registered address is changed), and our findings cannot be applied to such individuals.

It is important to note that the life expectancy estimates presented are the average across care-home residents and for individual residents the age at which death occurs will vary considerably. The data presented are reported by age and sex, but not adjusted for any other clinical conditions. This may not be a major issue for comorbidity, which is less predictive of mortality risk among those living in care-homes,^10^ but information on dependency or frailty,^29^ which is not systematically recorded in routine healthcare data, would likely have led to more precise estimates of life expectancy and may have caused us to over-estimate the YLL in those dying with COVID-19.

### Implications for practice, policy & future research

The most immediate priority we advocate is prioritising care-home residents and staff for vaccination. Our rationale is that by targeting 5% of the older adult population we can prevent around half of deaths and a quarter of years of life lost. Assuming vaccines are similarly efficacious across groups, vaccination strategies which specifically target care-homes are likely to have a larger impact on years of life lost than those which do not. Furthermore, these benefits may extend to those providing care to this vulnerable group as social care staff have experienced excess mortality during the pandemic.^30^

Even if vaccines rapidly bring the pandemic to an end, the observed two years life expectancy means that care-home residents will have had severely restricted access to family and friends for around half of their remaining lives. Balancing reducing COVID risk by restricting visiting with harm from social isolation has been one of the most difficult policy decisions during the pandemic. Future pandemic planning therefore needs to engage with residents and their families to discover how best this balance could be struck in the future. In communal settings, which are both people’s homes and places of work, where many residents have cognitive impairment, and where there are concerns about financial and even criminal consequences of these decisions, this process will be challenging and likely contentious. Nonetheless, accurate estimates of life expectancy will help inform such debates.

While not the main focus of this study, it is interesting to note the small trend before COVID-19 away from deaths in hospital.^13^ This likely reflects existing policy to encourage planned death in “homely” settings rather than hospitals, but it is unclear whether the larger drop in deaths in hospital during COVID-19 was appropriate or harmful and merits further exploration.

Beyond COVID-19, providing life expectancy estimates and historical comparisons for the care-home population is valuable. Individual and societal planning for care needs in older age relies on understanding the likely need for and costs of care to ensure needs are properly met. Understanding life expectancy may help clinicians, residents and their families make decisions about their health care, facilitating more informed discussions around their priorities and wishes.

## Conclusions

COVID-19 infection has led to the loss of substantial years of life in care-home residents aged 70 years and over in Scotland. Mortality impacts have been disproportionate on a vulnerable group. Life expectancy among those living in care-homes is consistently shorter than those of the same age and sex living elsewhere. This emphasises the need to prioritise the care of this group and consider how best to support their preferences and values and maximise their quality of life.

## Data Availability

All aggregated data (and analysis code) is available at https://github.com/ChronicDiseaseEpi/care_homes_covid_le. For access to the individual level anonymised data, apply to the Electronic Data Research and Innovation Service (eDRIS), information available from: www.isdscotland.org/Products-and-Services/eDRIS/

## Declarations

## Supplementary appendix – UPRN seeding of CHI

Public Health Scotland undertook work to seed the Community Health Index (CHI) database, of all General Practice registrations in Scotland, with a Unique Property Reference Number (UPRN), based on the individual’s registered address. This work was done in collaboration with the Improvement Service, utilising their Data Hub service. UPRN was derived from the Ordnance Survey Address Base

Address fields (3 address lines plus postcode) were extracted from the PHS CHI monthly download (dated 03-Aug-2020) from current records of persons still alive in Scotland or who died since 1st January 2020. This yielded 3,208,951 unique address strings relating to 5,828,951 people. Some minimal formatting and pre-processing was carried out in order to extract a Town entity from the address lines, trim leading spaces, format postcodes, and apply a series of rules to expand abbreviations at end of address lines (e.g. “RD” to “ROAD”, “DR” to “DRIVE” etc.). Files of unique address strings were processed at DataHub - https://datahub.scot/home/ - using the Non-residential UPRN seeding templates and programs. These returned 5 categories of output file with best matching UPRN attached to the input strings where applicable. The categories of output were EXCELLENT/GOOD/FAIR/MULTIPLE/NO_MATCH and the output is available from: https://www.scadr.ac.uk/sites/default/files/CURLreport2311%20-%20A%20guide%20to%20CHI-UPRN%20Residential%20Linkage.pdf

Additional manual work was undertaken to identify care-home addresses from the unmatched sample (441,550 unique addresses). Addresses were matched based on care-home name and partial postcode to known care-home addresses by a single reviewer, experienced in care-home address matching. Additional review was undertaken of addresses which had matched manually to identify care-homes in which there were no recorded residents or a mismatch of >10 registered places vs records in CHI. This work was done independently from the Institution Flag allocation. This manual work was necessary to maximise correct allocation of care-home addresses to UPRNs identifying care-homes.

## Supplementary figure – cumulative length of care home stay

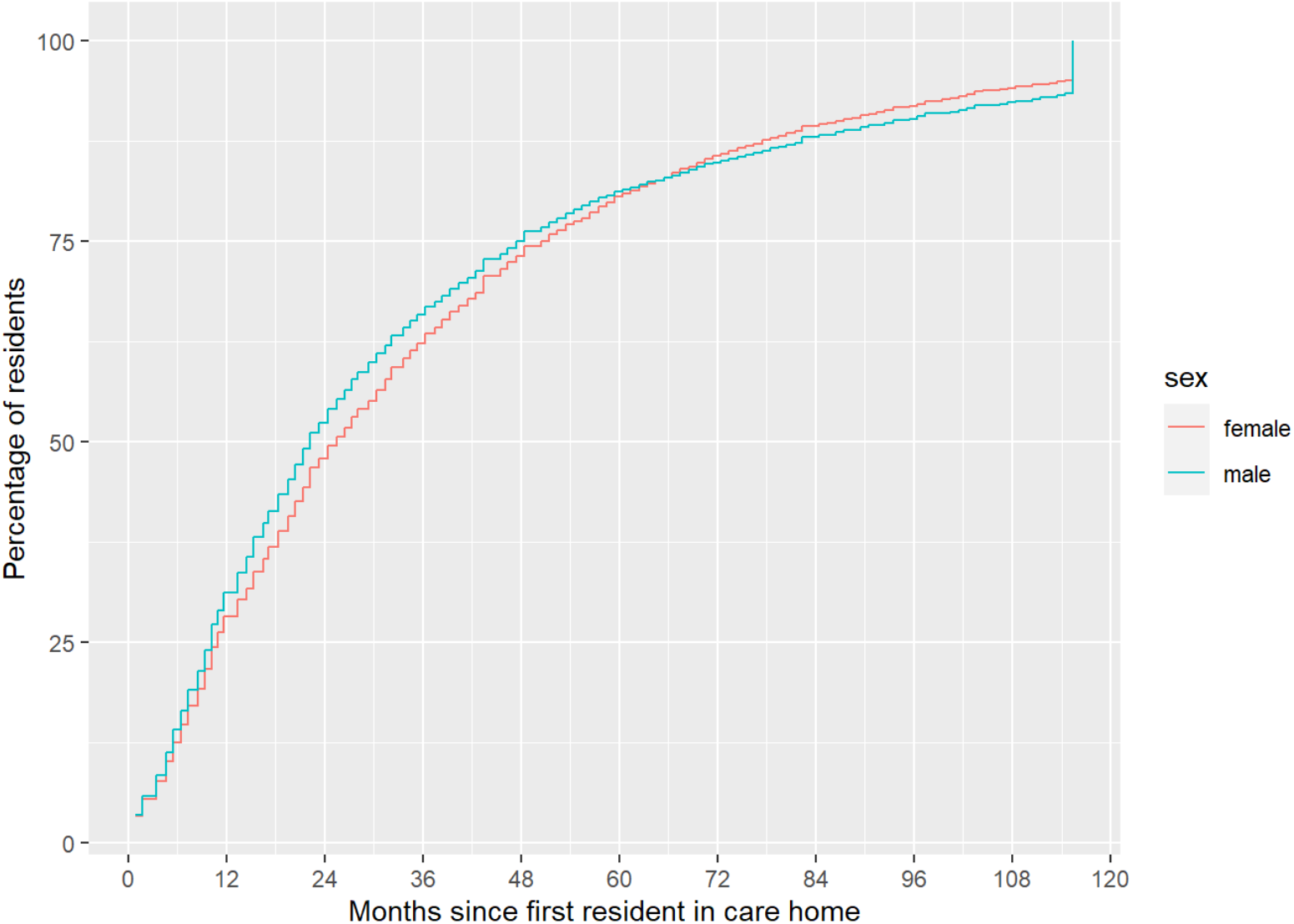
Red lines are rates in women and blue in men.

